# Colorectal peritoneal metastasis incidence and survival in the United States: A SEER retrospective cohort study

**DOI:** 10.1101/2025.03.02.25323124

**Authors:** Crystal Li, Aprill Park, Priyanka Ravi, Sumiao Pang, Magesh Sundaram, Jeremy L. Davis, Huang Chiao Huang, Benjamin D. Powers

## Abstract

**Background:** There are limited data on the incidence, distribution, and prognosis of colorectal peritoneal metastasis. However, some studies have suggested that colorectal peritoneal metastasis has a worse prognosis that other sites of metastasis. Therefore, this study assessed the epidemiology and prognosis of colorectal metastasis categories in the United States.

**Methods:** The incidence-based SEER database was used to identify patients with metastatic colorectal adenocarcinoma diagnosed from 2018-2021. Extent of disease and staging variables were used to determine sites of metastasis. Demographic and treatment variables were included as covariates. Descriptive statistics and survival analyses were conducted.

**Results:** The cohort consisted of 12,117 patients and distribution by staging category was M1a (46%), M1b (26%), M1c (19%), and uncategorized (9%). Among patients with M1c disease, 46.7% had peritoneal-only metastasis, comprising 9% of the entire cohort. The M1c cohort had a higher proportion of right-sided tumors (45.7%) and mucinous histology (10%) compared to M1a (33% and 2%, respectively) or M1b (30% and 2%, respectively). Median overall survival for M1a, M1b, and M1c was 20, 11, and 11 months, respectively (p<0.05). In adjusted analysis, the restricted mean survival time was 21.6, 17.1, and 17.5 months for M1a, M1b, and M1c cohorts, respectively and for M1c isolated and multisite metastasis was 20.3 and 14.7 months, respectively (p:<0.001).

**Conclusions:** Colorectal peritoneal metastasis accounted for approximately 20% of synchronous metastatic colorectal cancer cases. Although the absolute survival differences between M1b and M1c disease were negligible, there were significant survival differences between isolated M1c peritoneal-only disease compared M1c multisite (peritoneal and hematogenous) metastasis. These data demonstrate that colorectal peritoneal metastasis is not rare and the survival differences in the M1c subgroup have implications for clinical staging and suggest unique tumor biology.

## Introduction

Colorectal cancer is the fourth most common cancer diagnosed in the United States with an estimated 152,810 new cases in 2024.^1^ While the overall death rate has decreased, colorectal cancer remains the second-leading cause of cancer-related deaths.^1^ Approximately 22% of patients with colorectal cancer present with metastatic disease.^2^ However, up to 70% of patients may ultimately progress to metastatic disease or experience metastatic recurrence and most colorectal mortality is attributed to metastatic disease.^3^

Colorectal peritoneal metastasis has a particularly poor prognosis.^4, 5^ Clinical trial data have shown that colorectal peritoneal metastasis has a lower survival rate than liver or lung metastasis.^6^ However, few studies have analyzed the clinical outcomes of colorectal peritoneal metastasis. Previous reports examining colorectal peritoneal metastasis epidemiology predate the advent of fluoropyrimidine-based combination chemotherapy with oxaliplatin or irinotecan, have been single-center or randomized trial cohorts, or have evaluated trends using non-incident data.^6–12^ Additionally, progress has been limited by the lack of detailed, site-specific metastasis in national and population-based registries.

Despite the limited data showing a poor prognosis for colorectal peritoneal metastasis, the 8^th^ edition of the American Joint Committee on Cancer (AJCC) Cancer Staging Manual has introduced a new staging category, M1c, which denotes “metastasis to the peritoneal surface is identified alone or with other site or organ metastases.”^13^

Prior to the inclusion of the M1c category, peritoneal metastasis was not a distinct variable in the cancer data reported by the Surveillance, Epidemiology, and End Results (SEER) Program. Therefore, capitalizing on this change, data from the SEER cancer registry were used to assess the incidence, characteristics, and survival of metastatic colorectal cancer by AJCC 8^th^ edition staging categories and assess survival of the M1c cohort by isolated peritoneal disease and peritoneal plus extraperitoneal (hematogenous) sites of metastasis. These findings provide practicable, population-based estimates of colorectal metastasis incidence and survival and overcome challenges of limited external validity of clinical trial or single-center data.

## Materials and Methods

### Study Setting and Cohort

Using SEERStat statistical software version 8.4.4, the SEER 17 Research Plus cancer registry (November 2023 submission) was queried for patients with AJCC 8^th^ edition stage IV colorectal cancer. SEER collects cancer incidence and survival data from population registries, with SEER 17 covering over 26% of the United States population.^14^ The latest SEER registry version 22 includes 5 more registries and covers nearly 48% of the U.S. population but has limited field data no incidence data. Therefore, the SEER 17 dataset was utilized for this analysis. Because the data were de-identified, the study was deemed exempt from IRB review.

The study included patients over the age of 18 diagnosed with metastatic colorectal cancer (Supplemental Figure 1). *International Classification of Diseases for Oncology* codes C18.0 to C18.9 and C19.9 and C10.9 were included. Histologic type for carcinoma and adenocarcinoma were included (ICD-O-3 8000, 8010, 8020, 8140, 8480, 8481, and 8490). Appendiceal tumors, non-carcinoma or adenocarcinoma histology (e.g., neuroendocrine tumor and gastrointestinal tumor), patients diagnosed at autopsy or by death certificate and patients with more than two malignancies were excluded.

### Variable Selection

The predictor variable was AJCC Staging Manual 8^th^ Edition metastatic categories. These categories include M1a (metastasis to one site or organ, excluding peritoneal metastasis), M1b (metastasis to two or more sites or organs, excluding peritoneal metastasis), and M1c (peritoneal surface metastasis with or without another site or organ metastasis). Early-onset colorectal cancer was defined as patients with colorectal cancer under the age of 50.^15^ Covariates in the study were gender, race, ethnicity, primary tumor site (right colon C18.0-18.4, left colon C18.5-18.7, overlapping colon C18.8 and 19.9, and rectum C20.9), tumor grade, mucinous histology, surgery of the primary site, reason for no cancer directed surgery, and receipt of chemotherapy. To identify colorectal peritoneal metastasis with peritoneal only and peritoneal and additional metastatic sites, the M1c cohort was further characterized using site-specific metastatic variables. The M1c cohort with metastasis at diagnosis to distant lymph nodes, liver, lungs, and bone was defined as multisite (peritoneal and hematogenous) metastasis. The M1c cohort without metastasis at diagnosis to distant lymph nodes, liver, lungs, and bone was defined as isolated (peritoneal-only) metastasis. The primary outcome was overall survival (OS) and follow-up time was calculated form diagnosis until death or date of last contact.

### Statistical Analysis

For incidence analysis, rates were age-adjusted and presented per 100,000 of the US standard population with 95% confidence intervals.^16^ Patient characteristics were summarized using the median and range (25^th^ and 75^th^ percentiles) for continuous measures and frequencies and proportions for categorical measures. The association of metastatic category and continuous variables was assessed using Wilcoxon rank sum tests. For categorical variables Chi-squared tests or Fisher’s exact tests were used when the expected cell frequencies were less than 5. The reverse Kaplan-Meier estimator was used to calculate follow-up time.^17^ Kaplan-Meier survival estimates for OS were compared using the Log-rank test. Cox proportional hazard regression was used to fit unadjusted and adjusted (multivariable) models for metastatic staging category and covariates. The proportional hazards assumption was assessed with Schoenfeld residuals and time-varying covariates; however, the proportional hazard assumption was not met. Next, landmark models were fit for early and late events using interval-specific censoring, which also resulted in a violation of the proportional hazards assumption.^18^ Given these issues, the restricted mean survival time (RMST) approach was used as previously described.^19–21^ RMST is the area under the survival probability curve to a pre-specified time, which is described as τ (tau). The advantages of the RMST include avoiding the proportional hazards assumption, providing an absolute measure of effect size instead of a relative measure, and the ability to adjust for potential confounders.^20^ Due to the challenges with semi-parametric models, adjusted RMSTs were calculated and differences compared.^21^ For this analysis, τ was prespecified at 24 months and 46 months, the latter of which marked the last follow-up time.

RMST was reported with standard error. Due to the exploratory nature of this analysis p-values were not adjusted for multiple comparisons. All comparisons were two-sided and an association was considered statistically significant if *p* < 0.05. All analyses were performed with R version 4.4.2.

## Results

### Characteristics and Incidence of the Study Population

A total of 49,877 colorectal cases were identified of which, 12,117 cases (24%) met the inclusion criteria (Table 1). The median age at diagnosis of synchronous metastatic colorectal cancer was 65 years and females comprised 45% of the cohort. The distribution of M1 categories was M1a (46%), M1b (26%), M1c (19%), and uncategorized (9%). Among patients with M1c disease, 47% had isolated peritoneal metastasis, corresponding to 9% of the entire cohort, while 53% had multisite metastasis. Chemotherapy was received by 65% of the cohort.

**Table 1:**
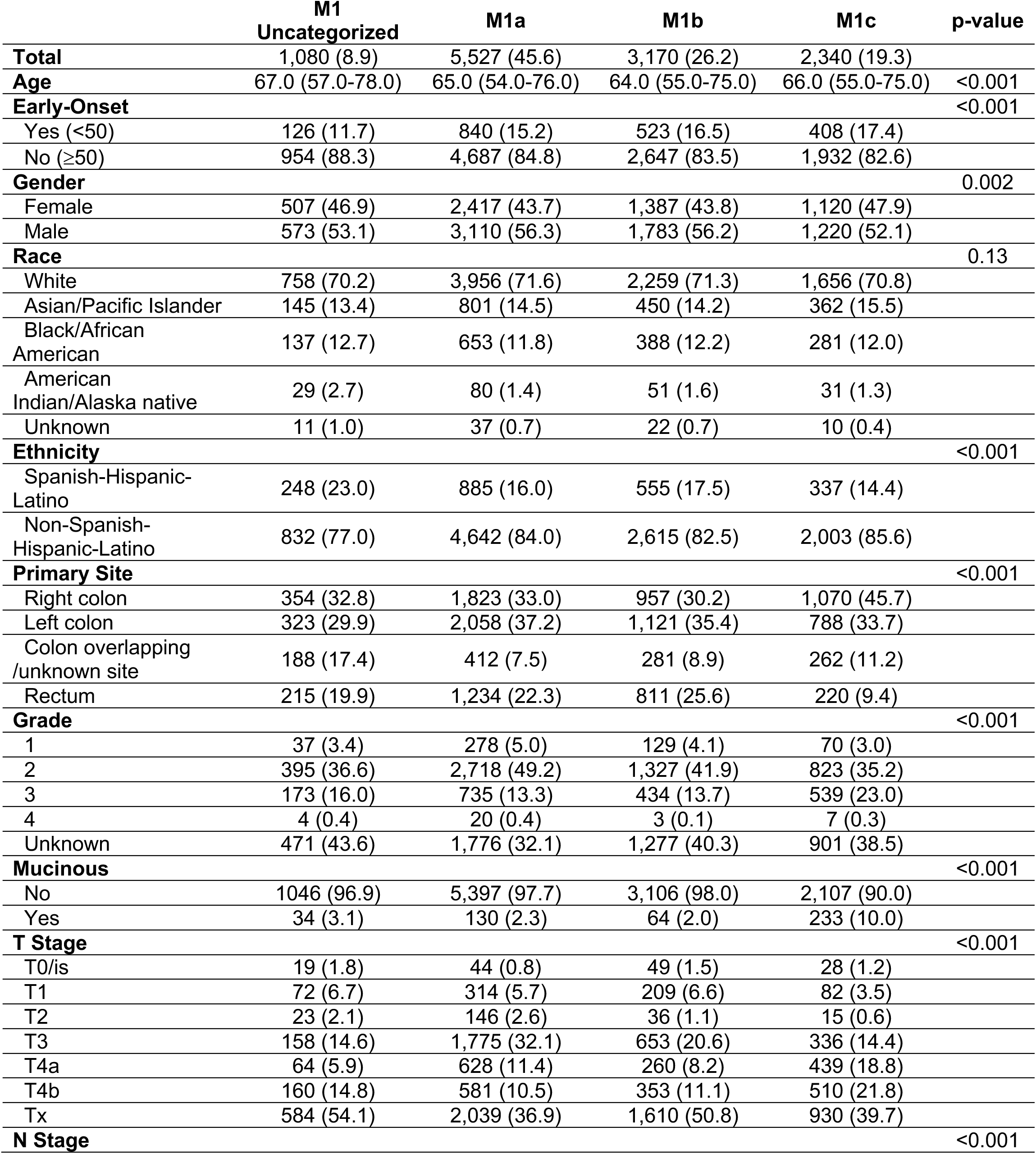
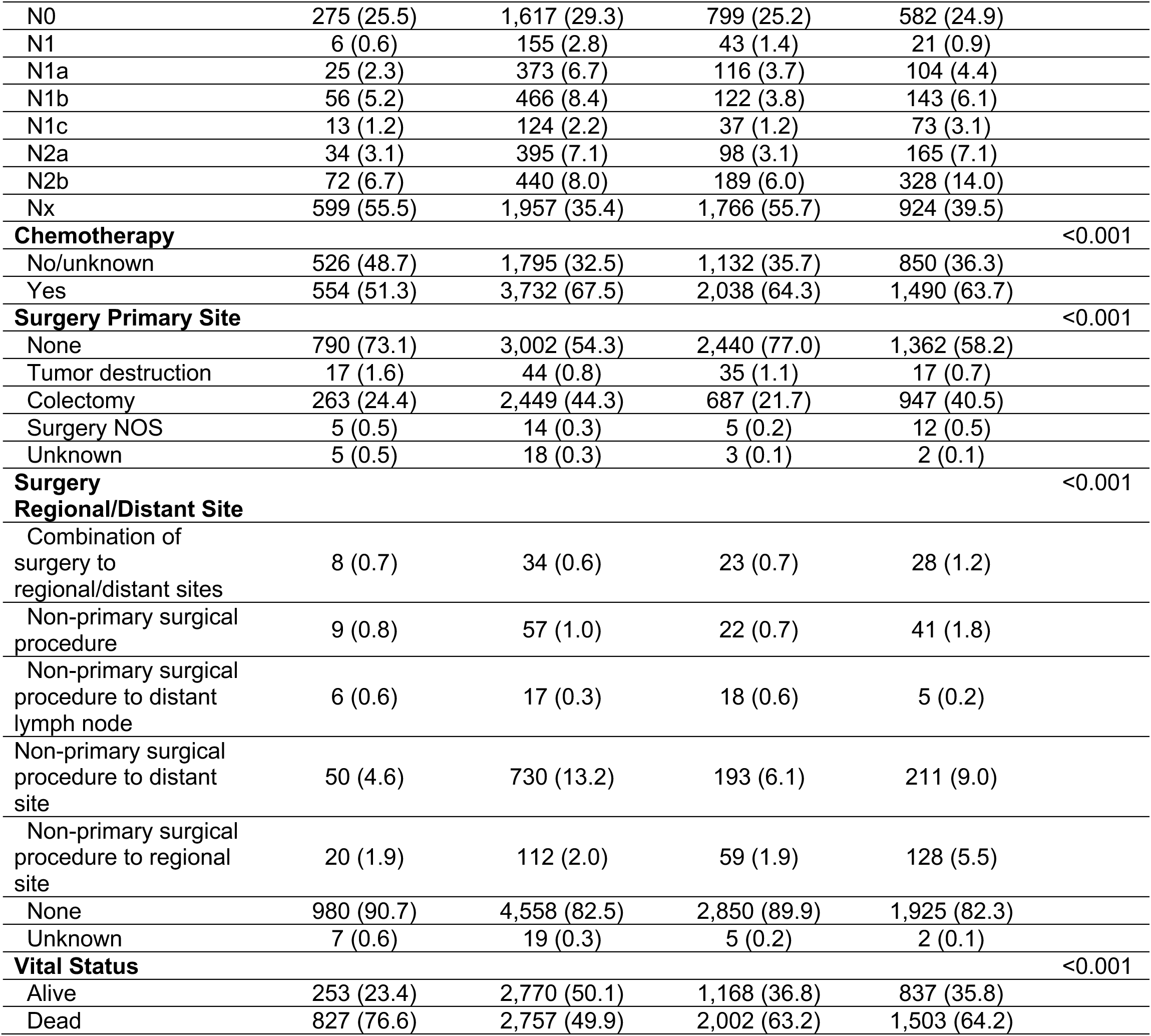
Characteristics of Colorectal Metastasis by Staging Category.

There were differences in the association of metastatic staging category and age, early-onset cancer, gender, ethnicity, primary site of disease, grade, mucinous histology, T and N stage, receipt of chemotherapy, receipt of surgery, and mortality (p<0.05). There were also differences in the association of metastatic staging category by isolated and multisite peritoneal disease and gender, ethnicity, primary site of disease, grade, mucinous histology, T and N stage, receipt of chemotherapy, receipt of surgery, and mortality (p<0.05; Supplemental Table 1).

Age-adjusted incidence per 100,000 persons by metastatic staging category is presented in Table 2. The incidence for the cohort was 7.1. By metastatic category, the M1a grouping had the highest incidence from 2018 to 2021 at 3.3, followed by the M1b (1.8) and M1c (1.3) groups. However, when stratified as by primary site (colon or rectal adenocarcinoma), there were differences in incidence by staging category. While colon metastasis incidence was similar for M1b (1.2) and M1c (1.1), rectal cancer incidence was 0.6 for M1b and 0.2 for M1c.

**Table 2:**
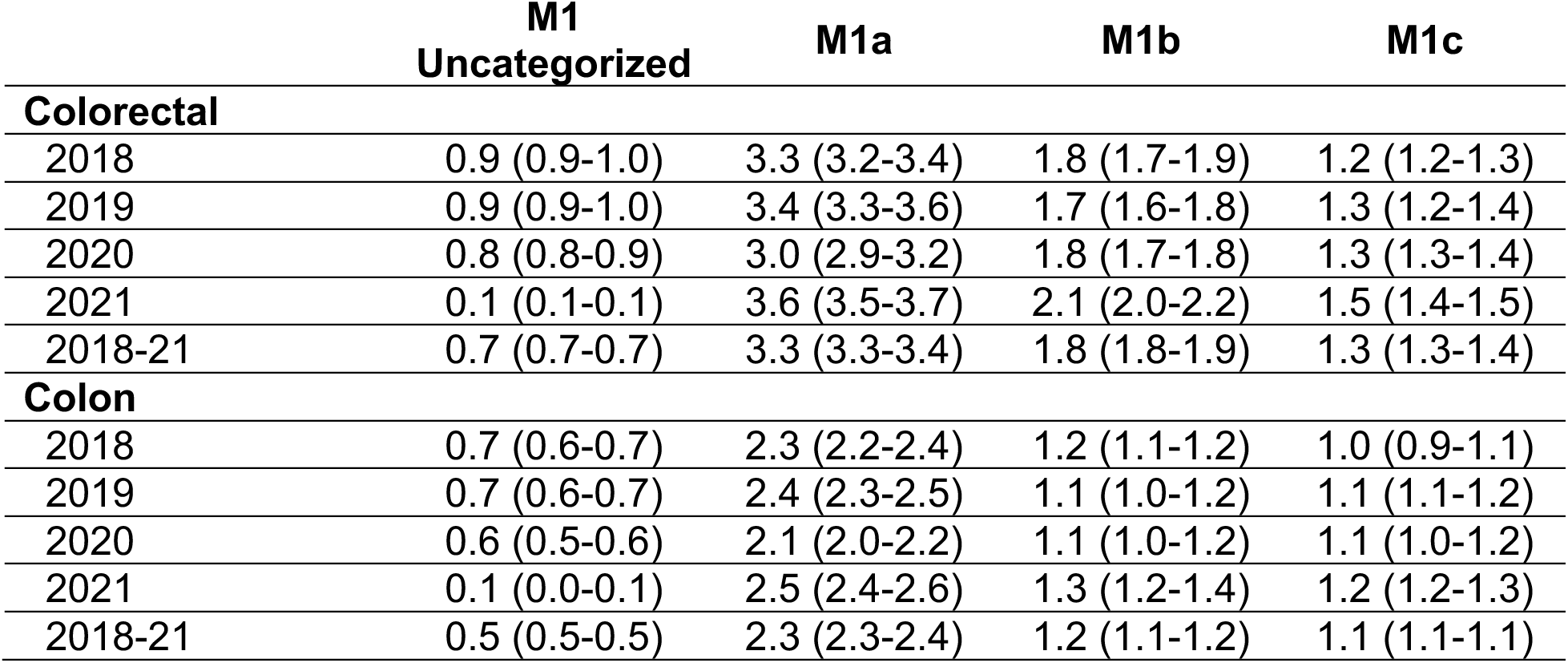
Age-Adjusted Incidence per 100,000 by Metastatic Colorectal Cancer Staging Category (95% Confidence Intervals)

|  | **M1**<br>**Uncategorized** | **M1a** | **M1b** | **M1c** |
| --- | --- | --- | --- | --- |
| **Colorectal** |  |  |  |  |
| 2018 | 0.9 (0.9-1.0) | 3.3 (3.2-3.4) | 1.8 (1.7-1.9) | 1.2 (1.2-1.3) |
| 2019 | 0.9 (0.9-1.0) | 3.4 (3.3-3.6) | 1.7 (1.6-1.8) | 1.3 (1.2-1.4) |
| 2020 | 0.8 (0.8-0.9) | 3.0 (2.9-3.2) | 1.8 (1.7-1.8) | 1.3 (1.3-1.4) |
| 2021 | 0.1 (0.1-0.1) | 3.6 (3.5-3.7) | 2.1 (2.0-2.2) | 1.5 (1.4-1.5) |
| 2018-21 | 0.7 (0.7-0.7) | 3.3 (3.3-3.4) | 1.8 (1.8-1.9) | 1.3 (1.3-1.4) |
| **Colon** |  |  |  |  |
| 2018 | 0.7 (0.6-0.7) | 2.3 (2.2-2.4) | 1.2 (1.1-1.2) | 1.0 (0.9-1.1) |
| 2019 | 0.7 (0.6-0.7) | 2.4 (2.3-2.5) | 1.1 (1.0-1.2) | 1.1 (1.1-1.2) |
| 2020 | 0.6 (0.5-0.6) | 2.1 (2.0-2.2) | 1.1 (1.0-1.2) | 1.1 (1.0-1.2) |
| 2021 | 0.1 (0.0-0.1) | 2.5 (2.4-2.6) | 1.3 (1.2-1.4) | 1.2 (1.2-1.3) |
| 2018-21 | 0.5 (0.5-0.5) | 2.3 (2.3-2.4) | 1.2 (1.1-1.2) | 1.1 (1.1-1.1) |

### Overall Survival

There were 7,089 deaths in the synchronous metastatic colorectal cancer cohort. The median follow-up time was 23 months and the median overall survival was 16 months. As shown in Table 3 and Figure 1, there was a difference in median overall survival by metastatic staging category (p<0.001). Median overall survival was 20 months for M1a, followed by 11 months for both M1b and M1c, respectively. For M1 uncategorized, the median overall survival was 8 months. There was a difference when M1c was assessed by isolated peritoneal disease or multisite metastasis, with a median survival of 13 and 8 months, respectively (p<0.001). For the cohort receiving chemotherapy, M1a had the longest median overall survival of 30 months. M1b and M1c had a similar median overall survival of 18 and 17 months, respectively. For M1c isolated peritoneal disease and multisite metastasis, the median overall survival was 19 and 15 months, respectively.

**Figure 1:**
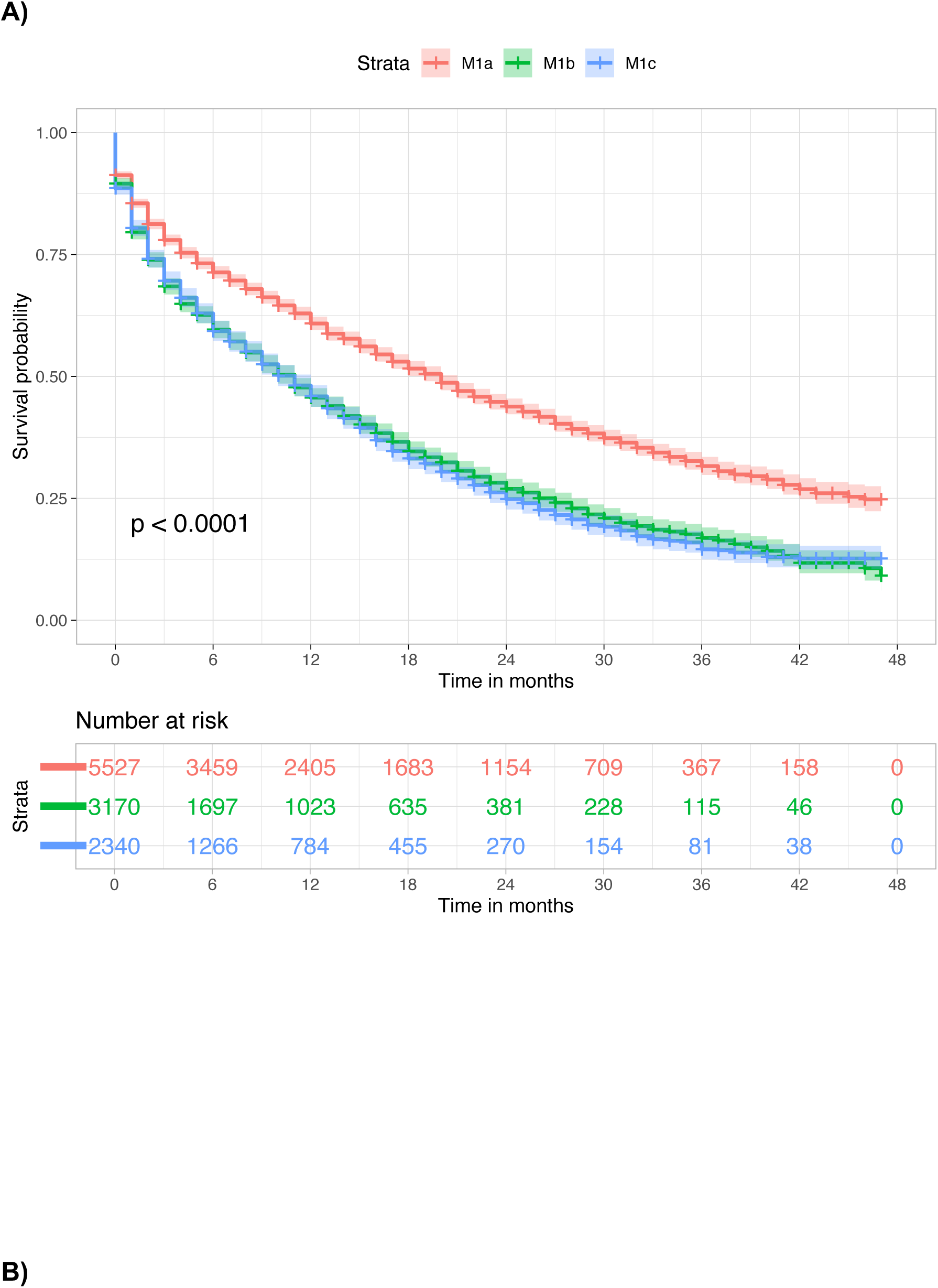
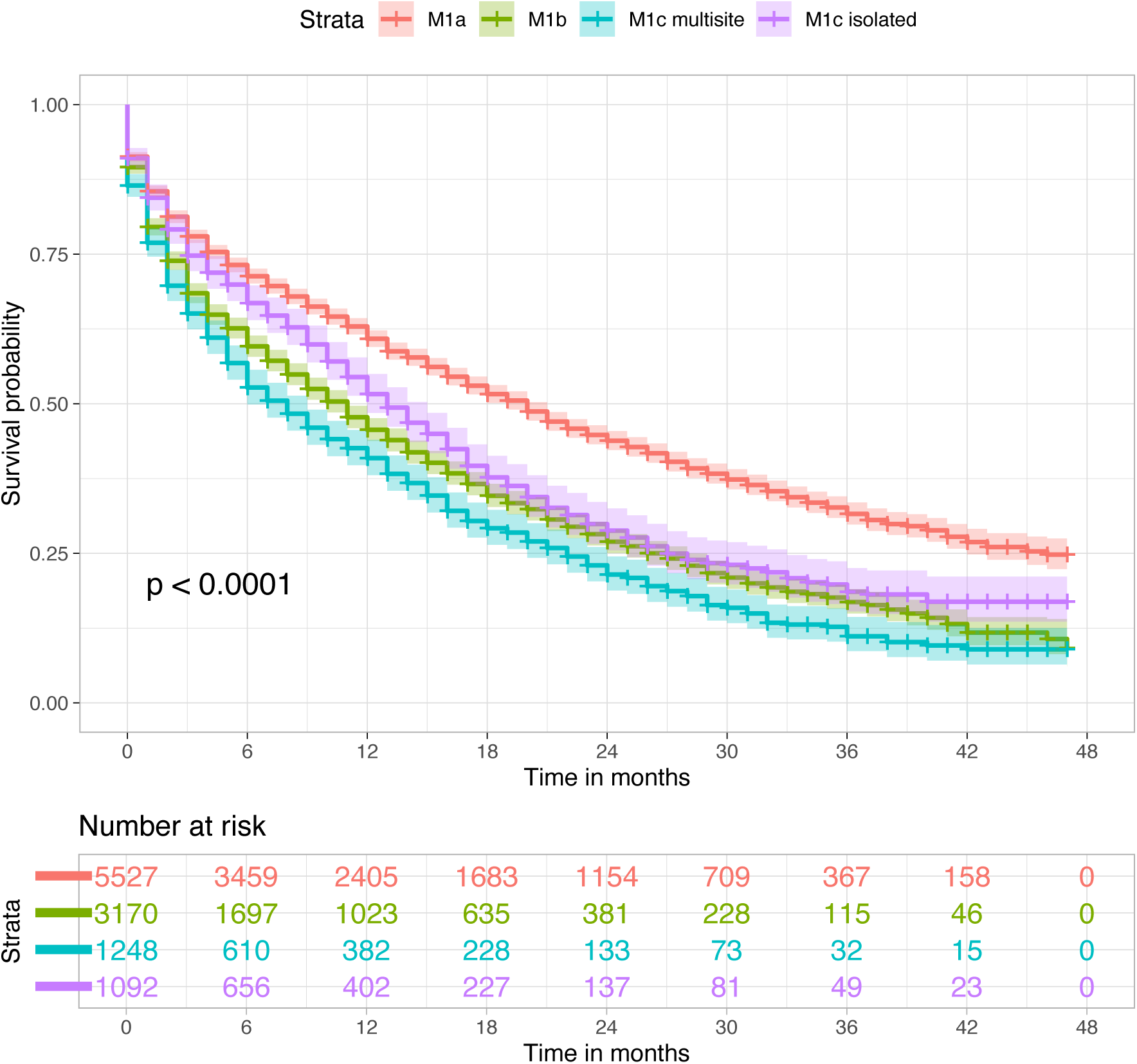
Kaplan Meier Overall Survival Curve of A) Colorectal Metastasis Staging Categories B) Colorectal Metastasis Staging Categories by M1c Isolated and Multisite Metastasis.

**Table 3:** A) Overall Survival of Colorectal Metastasis by Staging Category and by B) Receipt of Chemotherapy.

| **Total** | **N (events)** | **Median OS (95% CI)** |
| --- | --- | --- |
| M1 undefined | 1080 (827) | 8 (7-9) |
| M1a | 5527 (2757) | 20 (19-21) |
| M1b | 3170 (2002) | 11 (10-11) |
| M1c | 2340 (1503) | 11 (10-12) |
| M1c isolated | 1092 (640) | 13 (12-15) |
| M1c multisite | 1248 (863) | 8 (6-9) |

**B)**
| **Chemotherapy** | **N (events)** | **Median OS (95% CI)** |
| --- | --- | --- |
| M1 undefined | 554 (363) | 17 (15-19) |
| M1a | 3732 (1361) | 30 (28-32) |
| M1b | 2038 (1021) | 18 (17-19) |
| M1c | 1490 (801) | 17 (16-18) |
| M1c isolated | 679 (318) | 19 (17-21) |
| M1c multisite | 811 (483) | 15 (13-16) |
| **No/Unknown Chemotherapy** |  |  |
| M1 undefined | 526 (464) | 2 (1-2) |
| M1a | 1795 (1396) | 3 (2-3) |
| M1b | 1132 (981) | 1 (1-2) |
| M1c | 850 (702) | 2 (2-2) |
| M1c isolated | 413 (322) | 3 (2-4) |
| M1c multisite | 437 (380) | 1 (1-1) |

**Table 4:** Adjusted Restricted Mean Survival Time at A) 1 = 24 months B) τ = 46 months.

|  | **RMST, m (SE)** | **Difference in RMST, m (SE)**<br>**ref: M1a** | **Adjusted p**<br>**value** |
| --- | --- | --- | --- |
| **M1a** | 14.6 (0.13) | N/A | N/A |
| **M1b** | 12.1 (0.16) | 2.5 (0.21) | <0.001 |
| **M1c** | 12.4 (0.18) | 2.2 (0.22) | <0.001 |
| **M1c isolated** | 13.8 (0.27) | 0.8 (0.30) | 0.001 |
| **M1c multisite** | 10.9 (0.27) | 3.7 (0.30) | <0.001 |

**B)**
|  | **RMST, m (SE)** | **Difference in RMST, m (SE)**<br>**ref: M1a** | **Adjusted p**<br>**value** |
| --- | --- | --- | --- |
| **M1a** | 21.6 (0.26) | N/A | N/A |
| **M1b** | 17.1 (0.32) | 4.5 (0.41) | <0.001 |
| **M1c** | 17.5 (0.37) | 4.1 (0.45) | <0.001 |
| **M1c isolated** | 20.3 (0.55) | 1.3 (0.60) | 0.001 |
| **M1c multisite** | 14.7 (0.45) | 6.9 (0.54) | <0.001 |

Numbers are estimates with standard error (SE). Analyses are adjusted for age, gender, race, grade, sidedness, receipt of chemotherapy. RMST, restricted mean survival time; m, months.

After adjustment for potential confounders, RMST by staging category at 24 months of follow-up time was 14.6, 12.1, and 12.4 months for the M1a, M1b, and M1c cohorts, respectively (p<0.001). The RMST for M1c isolated metastasis to the peritoneum and M1c multisite metastasis was 13.8 and 10.9 months, respectively (p:<0.001). At a follow-up time of 46 months, the RMST was 21.6, 17.1, and 17.5 months for M1a, M1b, and M1c cohorts, respectively (p<0.001).

The RMST for M1c isolated and multisite metastasis was 20.3 and 14.7 months, respectively (p:<0.001). The greatest absolute RMST difference was between M1a and M1c multisite cohorts (6.9 months). There was negligible absolute RMST difference between the M1b and M1c cohorts (0.4 months). However, the difference between M1b disease and M1c multisite disease was 2.4 months and between M1b disease and M1c isolated disease was 2.8 months, respectively. Between isolated (20.3 month) and multisite (14.7 month) M1c disease, the absolute survival difference was 5.6 months.

## Discussion

To the authors’ knowledge, these findings represent one of the largest, incidence-based, studies of contemporarily treated colorectal peritoneal metastasis to date. The data show that colorectal peritoneal metastasis accounted for approximately 20% of incident cases of metastatic colorectal cancer with almost half of those being peritoneal-only disease. Furthermore, there was minimal overall survival difference between colorectal cancer M1b and M1c categories; however, there were significant differences in overall survival when the M1c cohort was categorized as isolated (peritoneal-only) metastasis and multisite (peritoneal and extraperitoneal) metastasis. This finding was consistent after adjustment; there was a significant difference in survival between M1c isolated and multisite disease. Collectively, these findings show that colorectal peritoneal metastasis is not uncommon and that within the M1c staging category, there are significant survival differences based on extraperitoneal metastasis.

Previous studies in Europe have reported epidemiologic data on synchronous colorectal peritoneal metastasis and the findings in this manuscript offer similar results. The proportion of colorectal peritoneal metastasis in two studies was 4.8% and 4.3%, respectively, which are similar to the finding herein of 4.7% (2,340/49,877).^8, 9^ Additionally, the proportion of isolated colorectal peritoneal metastasis was similar in this study (46.6%) when compared to prior reports.^9^ However, few studies have reported on the incidence of colorectal cancer by metastatic staging category. While the age-adjusted incidence for all colorectal cancer types in the United States is 35.8 per 100,000, this study evaluated metastatic colorectal adenocarcinoma cases only and excluded appendiceal tumors, showing an incidence of 7.1.^22^ These data using the current AJCC staging category provide a baseline for any future studies comparing trends in colorectal metastasis. An additional and important finding is that for colon metastasis, the incidence of M1b disease is similar to M1c disease, reinforcing that peritoneal metastasis is roughly as common a presentation as non-peritoneal multisite disease.

The M1c category for colorectal cancer was introduced in the 8^th^ edition AJCC staging manual based on data suggesting that “the prognosis for peritoneal disease is worse than that for visceral metastases to one or more solid organs.”^13^ Epidemiologic studies of colorectal peritoneal metastasis have found that isolated peritoneal disease may be associated with worse survival compared to isolated single metastasis in other sites, such as the liver or lung.^9, 23, 24^ However, the findings herein do not support a clinically meaningful difference in overall survival between the AJCC 8^th^ edition M1c classification, which does not distinguish between isolated and multisite peritoneal metastasis and the non-peritoneal metastases to one or more solid organs cohort (M1b). The median overall survival estimates for M1b and M1c were both 11 months and the adjusted survival difference was 0.4 months. Franko et al. analyzed randomized clinical trial data of metastatic colorectal cancer cohort that received systemic therapy using the Aide et Recherche en Canérologie Digestive (ARCAD) foundation database.^6^ For non-peritoneal multisite metastasis (M1b-equivalent), the median overall survival was 15.7 months. Within M1c disease, they also found significant survival heterogeneity by the presence of extraperitoneal disease; isolated peritoneal disease compared to peritoneal with additional metastasis showed an overall survival of 16.3 months and 12.6 months, respectively, with an absolute difference of 3.7 months. In a more generalizable cohort than clinical trial participants, the data in this study produced similar results; in adjusted survival, the absolute survival difference between isolated M1c and multisite M1c disease was 5.6 months. While treatment for colorectal peritoneal metastasis with systemic treatment and the use of cytoreductive surgery with or without heated intraperitoneal chemotherapy for select patients has been shown to improve survival, these findings reinforce the underlying prognostic differences based on extraperitoneal disease, which is a cornerstone of patient selection.^25–27^

Prior reports have suggested that colorectal peritoneal metastasis is a distinct metastatic pathway compared to liver and other visceral organ metastases.^28, 29^ Furthermore, molecular characterization has shown that over 80% of colorectal peritoneal metastasis falls within the Consensus Molecular Subtypes (CMS) 4 classification, or mesenchymal subtype.^30^ Nevertheless, as these data show, nearly half of patients with peritoneal disease may have concurrent hematogenous metastasis. The next challenge to improve patient selection and treatment outcomes is to merge knowledge of the clinical epidemiology of colorectal peritoneal metastasis with its molecular characteristics.^31^

There are several limitations to consider when interpreting these findings. First, this is a retrospective cohort study and, therefore, is subject to selection bias and missing data. However, the SEER program has been described as an authoritative source of information on cancer incidence and survival and covers 48% of the United States population.^32^ Additionally, while an effort was made to define isolated peritoneal metastasis, this study was not able to exclude less common sites of metastasis when constructing this variable, such as skin and neurological metastasis. However, these sites constitute only 4% of colorectal metastasis.^7^ Another limitation is that these data represent synchronous metastasis and recurrence data are not provided in SEER. Finally, while chemotherapy is provided as a treatment variable, surgery in SEER is only reported as regional and distant treatment without characterization of the intent or extent of surgery. Due to this, it was not included in adjusted models. Despite these drawbacks, the findings presented are prognostic, which is critical to informing the course of disease and patient counseling.^33^

## Conclusion

This large, incident-based population study showed that for colorectal metastasis, there is minimal survival difference for AJCC 8^th^ edition staging M1b and M1c cohorts. Additionally, peritoneal metastasis has heterogenous survival outcomes based on the presence of extraperitoneal disease. These findings have implications for prognosis and future staging guidelines. Linking epidemiologic outcomes to tumor biology may clarify the underlying mechanisms of these findings.

## Author contributions

Study conceptualization/design: CL, AP, PR, MS, SP, JLD, HCH, BDP;

Data curation: CL, AP, PR, BDP;

Formal analysis/investigation: CL, AP, PR, BDP;

Manuscript writing: CL, AP, PR, BDP;

Manuscript review/approval: CL, AP, PR, MS, SP, JLD, HCH, BDP;

## Conflict of interest statement

The authors declare no conflicts of interest

## Data availability statement

The data that support the findings of this study are openly available from the Surveillance, Epidemiology and End Results Program at https://seer.cancer.gov.

## Funding information

None

## Acknowledgements

None

---

**Supplemental Figure 1:**
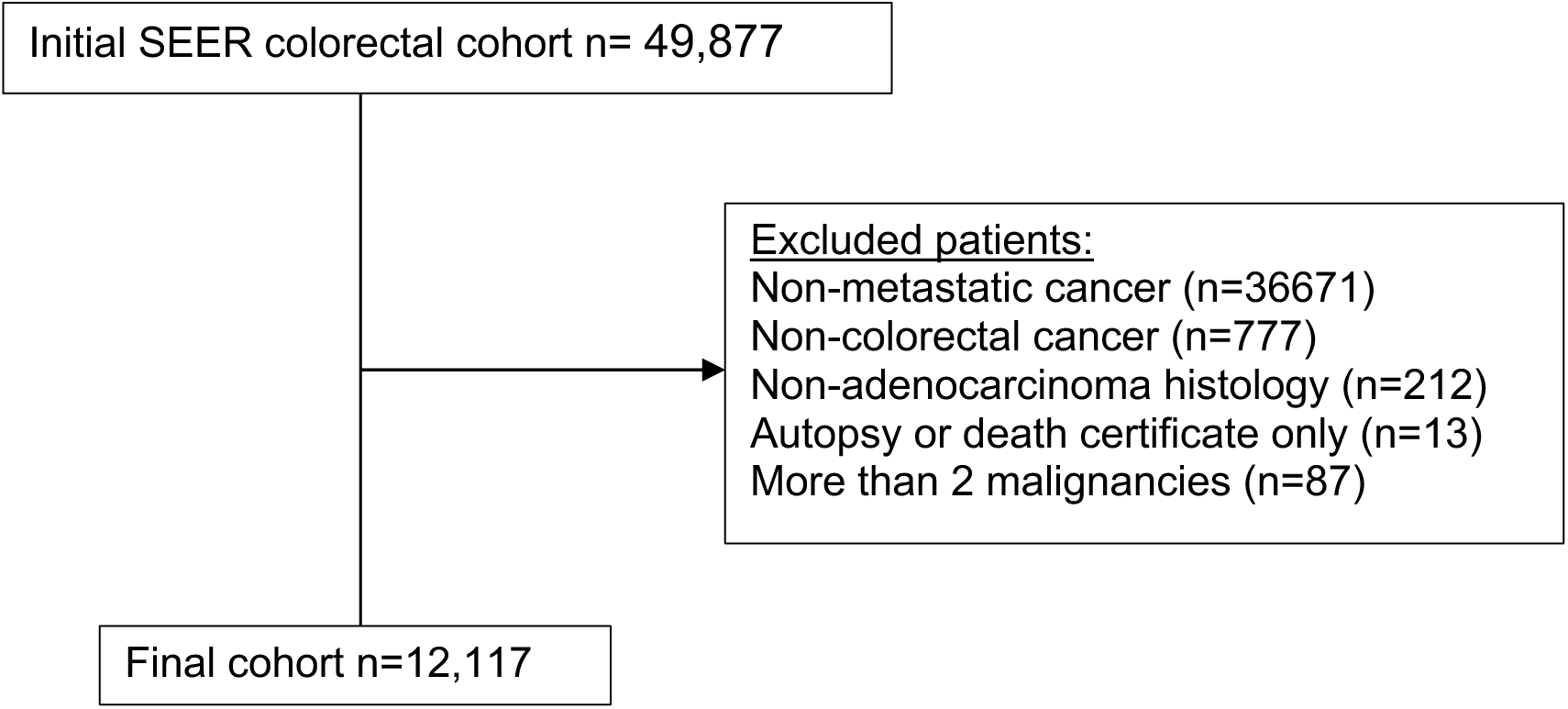
**Study population selection criteria**

**Supplemental Table 1:** Characteristics of Colorectal Metastasis Patients by M1c Distribution.

|  | **M1a** | **M1b** | **M1c Multisite** | **M1c Isolated** | **p-value** |
| --- | --- | --- | --- | --- | --- |
| **Total** | 5,527 (50.1) | 3,170 (28.7) | 1,248 (11.3) | 1,092 (9.9) |  |
| **Age** | 65.0 (54.0-76.0) | 64.0 (55.0-75.0) | 65.0 (54.0-75.0) | 66.0 (55.0-76.0) | 0.11 |
| **Early-Onset** |  |  |  |  | 0.078 |
| Yes (<50) | 840 (15.2) | 523 (16.5) | 216 (17.3) | 192 (17.6) |  |
| No (≥50) | 4,687 (84.8) | 2,647 (83.5) | 1,032 (82.7) | 900 (82.4) |  |
| **Gender** |  |  |  |  | <0.001 |
| Female | 2,417 (43.7) | 1,387 (43.8) | 573 (45.9) | 547 (50.1) |  |
| Male | 3,110 (56.3) | 1,783 (56.2) | 675 (54.1) | 545 (49.9) |  |
| **Race** |  |  |  |  | 0.58 |
| White | 3,956 (71.6) | 2,259 (71.3) | 875 (70.1) | 781 (71.5) |  |
| Asian/Pacific Islander | 801 (14.5) | 450 (14.2) | 183 (14.7) | 179 (16.4) |  |
| Black/African American | 653 (11.8) | 388 (12.2) | 167 (13.4) | 114 (10.4) |  |
| American Indian/Alaska native | 80 (1.4) | 51 (1.6) | 17 (1.4) | 14 (1.3) |  |
| Unknown | 37 (0.7) | 22 (0.7) | 6 (0.5) | 4 (0.4) |  |
| **Ethnicity** |  |  |  |  | 0.006 |
| Spanish-Hispanic-Latino | 4,642 (84.0) | 2,615 (82.5) | 1,083 (86.8) | 920 (84.2) |  |
| Non-Spanish-Hispanic-Latino | 885 (16.0) | 555 (17.5) | 165 (13.2) | 172 (15.8) |  |
| **Primary Site** |  |  |  |  |  |
| Right colon | 1,823 (33.0) | 957 (30.2) | 537 (43.0) | 533 (48.8) | <0.001 |
| Left colon | 2,058 (37.2) | 1,121 (35.4) | 433 (34.7) | 355 (32.5) |  |
| Colon overlapping /unknown site | 412 (7.5) | 281 (8.9) | 141 (11.3) | 121 (11.1) |  |
| Rectum | 1,234 (22.3) | 811 (25.6) | 137 (11.0) | 83 (7.6) |  |
| **Grade** |  |  |  |  | <0.001 |
| 1 | 245 (4.4) | 122 (3.8) | 26 (2.1) | 23 (2.1) |  |
| 2 | 2,158 (39.0) | 1,149 (36.2) | 325 (26.0) | 235 (21.5) |  |
| 3 | 489 (8.8) | 342 (10.8) | 140 (11.2) | 189 (17.3) |  |
| 4 | 8 (0.1) | 3 (0.1) | 1 (0.1) | 1 (0.1) |  |
| Unknown | 2,627 (47.5) | 1,554 (49.0) | 756 (60.6) | 644 (59.0) |  |
| **Mucinous** |  |  |  |  | <0.001 |
| No | 5,397 (97.6) | 3,106 (98.0) | 1,167 (93.5) | 940 (86.1) |  |
| Yes | 130 (2.4) | 64 (2.0) | 81 (6.5) | 152 (13.9) |  |
| **T Stage** |  |  |  |  | <0.001 |
| T0/is | 44 (0.8) | 49 (1.5) | 20 (1.6) | 8 (0.7) |  |
| T1 | 314 (5.7) | 209 (6.6) | 50 (4.0) | 32 (2.9) |  |
| T2 | 146 (2.6) | 36 (1.1) | 11 (0.9) | 4 (0.4) |  |
| T3 | 1,775 (32.1) | 653 (20.6) | 178 (14.3) | 158 (14.5) |  |
| T4a | 628 (11.4) | 260 (8.2) | 190 (15.2) | 249 (22.8) |  |
| T4b | 581 (10.5) | 353 (11.1) | 238 (19.1) | 272 (24.9) |  |
| Tx | 2,039 (36.9) | 1,610 (50.8) | 561 (45.0) | 369 (33.8) |  |
| **N Stage** |  |  |  |  | <0.001 |
| N0 | 1,617 (29.3) | 799 (25.2) | 293 (23.5) | 289 (26.5) |  |
| N1 | 155 (2.8) | 43 (1.4) | 14 (1.1) | 7 (0.6) |  |
| N1a | 373 (6.7) | 116 (3.7) | 42 (3.4) | 62 (5.7) |  |
| N1b | 466 (8.4) | 122 (3.8) | 56 (4.5) | 87 (8.0) |  |
| N1c | 124 (2.2) | 37 (1.2) | 32 (2.6) | 41 (3.8) |  |
| N2a | 395 (7.1) | 98 (3.1) | 80 (6.4) | 85 (7.8) |  |
| N2b | 440 (8.0) | 189 (6.0) | 145 (11.6) | 183 (16.8) |  |
| Nx | 1,957 (35.4) | 1,766 (55.7) | 586 (47.0) | 338 (31.0) |  |
| **Chemotherapy** |  |  |  |  | <0.001 |
| No/unknown | 1,795 (32.5) | 1,132 (35.7) | 437 (35.0) | 413 (37.8) |  |
| Yes | 3,732 (67.5) | 2,038 (64.3) | 811 (65.0) | 679 (62.2) |  |
| **Surgery Primary Site** |  |  |  |  | <0.001 |
| None | 3,002 (54.3) | 2,440 (77.0) | 830 (66.5) | 532 (48.7) |  |
| Tumor destruction | 44 (0.8) | 35 (1.1) | 8 (0.6) | 9 (0.8) |  |
| Colectomy | 2,449 (44.3) | 687 (21.7) | 403 (32.3) | 544 (49.8) |  |
| Surgery NOS | 14 (0.3) | 5 (0.2) | 6 (0.5) | 6 (0.5) |  |
| Unknown | 18 (0.3) | 3 (0.1) | 1 (0.1) | 1 (0.1) |  |
| **Surgery Regional/Distant Site** |  |  |  |  |  |
| Combination of surgery to regional/distant sites | 34 (0.6) | 23 (0.7) | 12 (1.0) | 16 (1.5) | <0.001 |
| Non-primary surgical procedure | 57 (1.0) | 22 (0.7) | 14 (1.1) | 27 (2.5) |  |
| Non-primary surgical procedure to distant lymph node | 17 (0.3) | 18 (0.6) | 4 (0.3) | 1 (0.1) |  |
| Non-primary surgical procedure to distant site | 730 (13.2) | 193 (6.1) | 98 (7.9) | 113 (10.3) |  |
| Non-primary surgical procedure to regional site | 112 (2.0) | 59 (1.9) | 60 (4.8) | 68 (6.2) |  |
| None | 4,558 (82.5) | 2,850 (89.9) | 1,059 (84.9) | 866 (79.3) |  |
| Unknown | 19 (0.3) | 5 (0.2) | 1 (0.1) | 1 (0.1) |  |
| **Vital Status** |  |  |  |  |  |
| Alive | 2,770 (50.1) | 1,168 (36.8) | 385 (30.8) | 452 (41.4) | <0.001 |
| Dead | 2,757 (49.9) | 2,002 (63.2) | 863 (69.2) | 640 (58.6) |  |

## Notes

### Competing Interest Statement

The authors have declared no competing interest.

### Funding Statement

This study did not receive any funding

